# Motor Imagery Ability and its Relationship with Psychosocial and Motor Variables in Patients with Chronic Shoulder Pain: A Cross-Sectional Study

**DOI:** 10.1101/2025.03.24.25324543

**Authors:** Mónica Grande-Alonso, Nerea Molina-Hernández, Bárbara Mira-Valverde, Celia Vidal-Quevedo, Carlos Forner-Alvarez, Ferran Cuenca-Martínez

## Abstract

**Objective:** To assess the ability to generate both kinesthetic and visual motor imagery in participants with chronic shoulder pain, compared with asymptomatic subjects. To assess the influence of psychosocial and motor variables in the motor imagery process.

**Methods:** A cross-sectional observational study was conducted with a non-probability sample of 30 participants, divided into 15 patients with chronic shoulder pain and 15 asymptomatic subjects. Participants completed socio-demographic questionnaires, followed by assessment of mental imagery and mental chronometry using the MIQ-R questionnaire, as well as range of motion and grip strength.

**Results:** Our results indicated that patients with chronic shoulder pain had difficulty generating kinesthetic and visual MI (p<0.05; d>0.80) and also took a longer time to imagine them (p<0.05; d>-0.80). A moderate-positive association was found between the levels of CPSS and the kinesthetic motor imagery ability (r=0.54, p=0.035), explaining 24.5% of the variance. In addition, a moderate-positive association was also found between the strength of the affected upper limb with the total score of MIQ-R (r=0.56, p=0.037), explaining 25.7% of the variance.

**Conclusions:** Based on the obtained results, it seems that patients with chronic shoulder pain have greater difficulty with generating both kinesthetic and visual motor images compared to asymptomatic subjects, and they also needed more time to perform the mental tasks. It was found that psychosocial state and motor condition may influence the imagining ability of patients with chronic shoulder pain. Further research is needed to improve our understanding and to be able to establish clinical implications.

## Introduction

Shoulder pain is considered the third most prevalent musculoskeletal problem with an average annual incidence of 37.8 per 1000 persons (Britt et al., 2015; Lucas et al., 2022). The one-year prevalence of shoulder pain has been reported as high as 55%. Moreover, shoulder pain is characterized by a tendency towards chronification, with around 50% of patients suffering from shoulder pain for at least six months (Kuijpers et al., 2004). It has been observed that patients suffering from chronic shoulder pain (CSP) show a decrease in range of motion, a direct impairment of quality of life, as well as an alteration of various psychological factors and, of course, impairment of other functional variables (Badcock et al., 2002; Hill et al., 2010).

Some evidence points to central sensitization (CS) as the potential cause of neural plasticity and central rearrangement in individuals with CSP, as well as a potential contributing factor to their recurrent complaints (Nijs et al., 2016). According to available data, individuals who report CSP related with CS, may also have sensorimotor and psychological disorders that play a role in the maintenance of their symptoms (Bilika et al., 2002). Furthermore, it has also been demonstrated that psychological variables, in addition to the presence of pain, can have a direct impact on motor planning and functionality (Hanakawa et al., 2008; Osumi et al., 2021; Summers et al., 2020). This can result in alterations in motor control and muscle activation, as well as a decrease in position sensing and stimulus perception (Gwilym et al., 2011; Struyf et al., 2015).

In addition to the aforementioned alterations that may exist in patients with CSP, it is possible that there are other alterations in these patients that have not yet been investigated, and that are present in other chronic pain conditions, such as alterations in the ability to generate motor imaging (La Touche et al., 2019; Moseley et al., 2008). Motor imagery (MI) can be described as a dynamic state in which a person mentally represents a specific action (Decety, 1996). Interestingly, there is an overlap between some structures that may be altered in a CSP process and are involved in the MI process, such as the motor cortex and the prefrontal cortex (Conboy et al., 2021; Ngomo et al., 2015).

Thus, both the fact that alterations in MI have been seen to exist in other chronic pain conditions and the overlap between structures affected in CSP and those involved in MI may make it more likely that alterations in MI exist in patients with CSP.

Thereby, the main aim of the present study was to assess the ability to generate both kinesthetic and visual MI in patients with CSP, compared with asymptomatic subjects. In addition, the secondary aim was to assess the influence of psychosocial and motor variables in the MI process.

## Methods

### Study design

A cross-sectional observational study was conducted between December 2022 and July 2023, in accordance with the criteria “Strengthening the Reporting of Observational Studies in Epideiomolgy” (STROBE) (Von Elm et al., 2008). This study was approved by the ethics committee of University La Salle (CSEULS-PI-031/2022). All participants who took part in the study received an information sheet and signed and dated the informed consent form in compliance with the declaration of Helsinki.

### Participants

A total of 30 participants were recruited, of which 15 were patients with CSP and 15 were asymptomatic subjects (AS). Patients with CSP were recruited from the La Salle Functional Rehabilitation Institute. The sample of AS was recruited from the local community via telephone or verbal communication. Participants were recruited between December 2022 and July 2023.

#### Inclusion criteria

The selection criteria for the AS were (a) men and women aged 18-65 years; (b) presenting with a primary complaint of unilateral shoulder pain with or without irradiation to the upper limb of more than 3 months’ duration; (c) with pain intensity greater than or equal to 3 points on the visual analogue scale (VAS) when performing any type of movement; (d) presence of pain on performance of shoulder abduction or external rotation; (e) presentation of minimal or no pain intensity at rest; and (f) pain provocation on at least three of the following tests: Hawkins-Kennedy, Neer’s, Jobe’s and on at least one of the following three: painful arc, dropped arm test and Gerber’s test.

The selection criteria for the CSP group were a) subjects (male and female) aged between 18 and 65 years not suffering from shoulder pain at the time of measurement or three months prior to the time of assessment; b) participants without the presence of systemic, neurological, cognitive or psychological disease.

#### Exclusion criteria

The exclusion criteria were: (a) presentation of history of previous shoulder surgery; (b) presence of signs of radiculopathy pain such as motor weakness, hyporeflexia or sensory disturbances; (c) presentation of diagnosed systemic diseases such as diabetes, fibromyalgia, rheumatoid arthritis, lupus and/or neoplasia; (d) presentation of pain of traumatic origin and/or previous shoulder fracture; e) external rotation of less than 45° or less than 50% relative to the contralateral shoulder (measured in a seated position, with 0° of shoulder abduction (R0 position) to rule out the presence of pathology compatible with frozen shoulder syndrome; and f) glenohumeral instability, including previous shoulder dislocation/subluxation. Positive sulcus sign and/or positive test (drawer test and/or apprehension test) for anterior and/or posterior instability.

### Procedure

After giving written consent to participate in this research, all recruited participants received a socio-demographic questionnaire to be completed on the day of the assessment (gender, date of birth, marital status and educational level, among others). Each participant then completed a series of self-report measures through questionnaires of psychological and disability variables. Next, each participant completed the Movement Imagery Questionnaire-Revised (MIQ-R) with the assessing physiotherapist, also assessing the time spent performing the tasks in this questionnaire. Finally, the assessing physiotherapist was in charge of measuring the range of movement (ROM) of flexion and abduction of the shoulder bilaterally and the grip strength of both hands.

### Variables

#### Primary variable

##### Visual and Kinesthetic Motor Imagery Ability

MIQ-R is an 8-item self-report inventory with adequate internal consistency (Cronbach’s alpha coefficients ranging above 0.84 for the total scale, 0.80 for the visual subscale, and 0.84 for the kinesthetic subscale). It was used to assess visual and kinesthetic motor imagery ability. Four different movements are including in this test, and the inventory is composed of 4 visual and 4 kinesthetic items. For each item, patients read a description of the movement. They then physically performed the movement before performing the mental task, which was to imagine the movement visually or kinesthetically. A score between 1 and 7 is assigned, with 1 representing difficulty in picturing the motor image or difficulty in feeling the movement previously made, and 7 representing the maximum ease. The time to perform each item also was evaluated (Campos A. & González MA., 2010).

##### Imagery-Requested Time

An imagery-requested time evaluation was also used to measure the patients motor imagery ability with a stopwatch. The time recorded corresponded to the interval between the command to start the task and the moment the task had been concluded (Williams et al., 2015).

#### Secondary variables

##### Pain intensity

The self-reported pain intensity in the lumbar area was assessed using the numerical pain scale. In this scale, a score of 0 indicates “no pain”, and a score of 10 indicates “the maximum pain intensity possible”. (Sendlbeck et al., 2015) It has been shown to have good validity (r=0.94, P<.001)

##### Flexión and abduction ROM

For the assessment of ROM, a validated mobile app (ICC=0.89) was used (Werner et al., 2014). Shoulder flexion and abduction movements were assessed on both sides. For this purpose, the mobile phone was placed on the anterior aspect of the proximal third of the humerus to measure the degrees of movement during abduction and on the lateral aspect of the proximal third of the humerus to measure the degrees of movement during shoulder flexion.

##### Manual grip strength

Bilateral manual grip strength was assessed asynchronously on each side using a dynamometer (ICC = 0.902 (0.789-0.954) to 0.990 (0.978-0.995), with the subject in a seated position, back against the backrest, both feet on the floor, arm resting on an armrest so that the elbow was in 90 degrees of flexion.(Morin et al., 2023) The procedure was repeated 2 times on each side, leaving a washout period of at least 1 minute between measurements. (Sánchez Torralvo et al., 2018)

##### Shoulder disability

For the assessment of disability induced by shoulder pain, the “Disabilities of the Arm, Shoulder and Hand” questionnaire was used in its reduced version: Quick DASH (17-Cronbach = 0.96). (Hervás et al., 2006) This questionnaire consists of 11 items in Likert format with 5 response options, ranging from “no difficulty” to “I cannot perform”. The higher the score, the lower the participant’s ability to perform activities of daily living. Scoring ranges from 11 to 55. Each item is scored from 1 to 5, with 1 point being no difficulty, 2 points being mild difficulty, 3 points being moderate difficulty, 4 points being severe difficulty and 5 points being I cannot perform. (Werner et al., 2014)

##### Anxiety and Depression

The Hospital Anxiety and Depression Scale (HADS) was used to assess traits related to anxiety and depression (test-retest reliability = 0.84-0.85 from 0 to 2 weeks, from 0.73 to 0.76 from 2 to 6 weeks, and more than 6 weeks was 0.70). (Herrero et al., 2003) This scale has a total of 14 items, divided into 2 subscales of 7 items, one for the assessment of traits related to anxiety and the other subscale for the assessment of traits related to depression. It is a Likert scale, in which each item has 4 response options. In relation to psychometric properties, Herrmann’s test-retest reliability was excellent, 0.84-0.85 from 0 to 2 weeks, 0.73-0.76 from 2 to 6 weeks, and over 6 weeks was 0.70. (Herrmann, 1997)

##### Kinesiophobia

For the assessment of kinesiophobia, the TAMPA Kinesiophobia Scale (TSK-11) was used, which consists of 11 items, in Likert format, with 4 response options ranging from 1 to 4, “strongly agree” to “strongly disagree”, respectively. To obtain the total scale score, the scores for each item are added together. The score ranges from 11 to 44 points, for each item the score ranges from 1 to 4, with 1 being strongly disagree, 2 somewhat disagree, 3 somewhat agree and 4 strongly agree. (Gómez-Pérez et al., 2011) Regarding psychometric properties, all versions of the questionnaire except TSK-4 show good to excellent test-retest reliability (intraclass correlation coefficient 0.77 to 0.99) and good internal consistency (= 0.68 to 0.91). (Dupuis et al., 2023)

##### Chronic pain self-efficacy

To assess self-efficacy, the Chronic Pain Self-Efficacy Questionnaire (CPSS) was used, which consists of 19 items, each of which determines the ability to perform a certain task, scored from “0 - I think I am totally incapable” to “10 - I think I am totally capable”. The questionnaire consists of 3 subscales: coping, function and pain. A higher score indicates better self-efficacy in pain management. (Martín-Aragón et al., 1999) The CPSS presented a reliability of 0.88, 0.87 and 0.90 for the subscales of Pain Control, Physical Functioning and Symptom Coping subscale, respectively. (Anderson et al., 1995)

##### Pain catastrophizing

Pain catastrophizing was assessed using the self-reported Pain Catastrophizing Scale (PCS), which consists of 13 items, in Likert format with 5 response options. It assesses 3 subscales: rumination, magnification and hopelessness. The total score ranges from 0 to 52 points, for each item the score varies between 0 and 4, with 0 being not at all, 1 a little, 2 moderately, 3 a lot and 4 all the time. (García Campayo et al., 2008) This instrument has been shown to be valid and reliable (Cronbach’s α, 0.79; ICC, 0.84).

##### Physical activity

Physical activity was assessed using the International Physical Activity Questionnaire (IPAQ). (Gauthier et al., 2009) This questionnaire consists of 7 items assessing the amount of physical activity performed in intense, moderate or walking. As well as the amount of time spent sitting.

By means of a formula, the minutes of intense activity are multiplied by 8 and by the total number of days per week in which this activity is performed. The same procedure is done for moderate activity by multiplying it by 4 and for walking activity by multiplying it by 3.3. Finally, the three values are added together to obtain a total number expressed in METs, which allows the subjects to be classified according to the amount of physical activity performed. Reliability analysis showed Spearman correlation coefficients between 0.96 and 0.46, but most of them were around 0.8 indicating good reliability. For the short version of the IPAQ, 75% of the correlation coefficients observed were above 0.65 with ranges between 0.88 and 0.32. (Gauthier et al., 2009)

### Sample Size Calculation

The sample size calculation is considered as a power calculation to detect between-group differences in the primary outcome measure (MI ability in the present study). To calculate the sample size, we used the results obtained from a previous observational study (Matesanz-García et al., 2023). The study conducted by Matesanz-García et al. (2023) found between-group statistically significant differences in the MI ability with a large effect size in a pilot study (d=1.6). They performed the sample size calculation with the program G*Power 3.1.7 for Windows (G*Power from University of Dusseldorf, Germany) (Faul et al., 2007) considering a statistical power of 90% with an α error level probability of 0.05 obtaining a total sample size of 30. Therefore, due to the similarity of our study with theirs, we also used the same sample size that they reported (n=30, 15 per group).

### Data analysis

The Statistical Package for Social Sciences (SPSS 25, SPSS, Chicago, IL) was used. Descriptive statistics showed data for continuous variables, in the form of means and standard deviations (SD), with 95% confidence interval (CI) and frequency relative to percentage (n (%)). The Chi-square test was used to compare the difference between nominal variables. Since each group consisted of less than 30 participants, normality tests were performed. With regard the main variables, Student’s t-test for independent samples was used as a statistical test to compare continuous variables between groups. The effect size (Cohen’s d) was then calculated to compare the study variables. According to Cohen’s method, the effect was considered small (0.20-0.49), medium (0.50-0.79) or large (>0.8) (Cohen, 1988). For the second objective, we ran Pearson’s correlation coefficient. A Pearson correlation coefficient >0.60 indicated a strong correlation, a coefficient between 0.30 and 0.60 indicated a moderate correlation and a coefficient <0.30 indicated a low correlation (Hinkle et al., 1990). A single linear regression analysis was performed to estimate the strength of the associations between the ability to generate both kinesthetic and visual motor images. Psychosocial and motor variables were used as predictors. The strength of the association was examined using regression coefficients (β), p-values, and adjusted R^2^. Standardized beta coefficients were reported for each predictor variable included in the final reduced models to allow for a direct comparison between the predictor variables in the regression model and the criterion variable being studied. For the data analysis, we used a CI of 95%, considering all those values that had a p-value of less than 0.05 to be statistically significant.

## Results

The total study sample consisted of 30 participants who were divided into two groups (15 patients with CSP, and 15 AS) who met the inclusion criteria. The **Table 1** showed the sociodemographic and participants characteristics data. The normality test found no statistical differences showing a normal distribution of the sample (p>0.05).

**Table 1.** Socio-demographic and participants characteristics.

| **Variables** | **CSP (n=15)** | **AS (n=15)** | **p-value** |
| --- | --- | --- | --- |
| **Age** | 50.8 ± 10.6 | 44.5 ± 16.0 | 0.21 |
| **Female** | 11 (73.3) | 6 (40) | 0.07 |
| **BMI** | 27.6 ± 5.0 | 25.7 ± 3.5 | 0.23 |
| **IPAQ** | 1763 ± 1362 | 2269 ± 1453 | 0.33 |
| **Pain intensity (now)** | 5.5 ± 1.7 | - | - |
| **Pain intensity (last week)** | 5.3 ± 1.4 | - | - |
| **Pain frequency (d/m)** | 25.6 ± 6.3 | - | - |
| **Chronicity (months)** | 25.8 ± 28.1 | - | - |
| **Affected upper limb** |  | - | - |
| **Left** | 8 (53.3) |  |  |
| **Right** | 7 (46.7) |  |  |
| **Dominant hand** |  |  |  |
| **Left** | 1 (6.7) | 0 (0) | 0.31 |
| **Right** | 14 (93.3) | 15 (100) |  |

*Notes: Data are presented as mean and standard deviation or n (%). CSP: Chronic shoulder pain; AS: Asymptomatic subjects; d/m: day per month; BMI: Body mass index; IPAQ: International physical activity questionnaire.*

### Motor imagery ability

The comparison analysis showed significant between-groups differences in the kinesthetic MIQ-R subscale score, with a large effect size (mean differences (MD)=6.2 (95% CI 3.5 to 8.8), t(28)=4.78, p<0.001, d=1.76), and also in the visual MIQ-R subscale score, with a large effect size (MD=4.7 (95% CI 2.6 to 6.8), t(28)=4.78, p<0.001, d=1.72). Finally, the results showed statistically significant between-groups differences in the total score of MIQ-R, also with a large effect size (MD=10.5 (95% CI 6.5 to 14.6), t (28)=5.4, p<0.001, d=1.96). These results found that patients with CSP showed a poorer MI ability compared to AS **(Table 2)**.

**Table 2.** Differences between patients and asymptomatic subjects in motor imagery ability.

| **Variables** | **CSP (n=15)** | **AS (n=15)** | **p-value** |
| --- | --- | --- | --- |
| **MIQ-R (K)** | 20.3 ± 4.6 | 26.5 ± 1.9 | <0.001** |
| **MIQ-R (V)** | 21.7 ± 3.5 | 26.4 ± 1.6 | <0.001** |
| **MIQ-R (Total)** | 42.2 ± 6.9 | 53.0 ± 2.9 | <0.001** |
| **MIQ-R (K) Time** | 19.6 ± 4.9 | 13.1 ± 3.6 | <0.001** |
| **MIQ-R (V) Time** | 16.9 ± 4.4 | 13.0 ± 3.1 | 0.009* |
| **MIQ-T (Total) Time** | 36.5 ± 8.7 | 26.3 ± 6.7 | 0.01* |

*Notes: \*p<0.05; \*\*p<0.001. MIQ-R: Movement Imagery Questionnaire-revised version; K: Kinesthetic subscale; V: Visual subscale; CSP: Chronic shoulder pain; AS: Asymptomatic subjects.*

With regard the mental chronometry, the comparison analysis showed significant between-groups differences in the kinesthetic MIQ-R subscale, with a large effect size (MD=−6.54 sec (95% CI −9.7 to −3.3), t(28)=4.15, p<0.001, d=−1.53), and also in the visual MIQ-R subscale, with a large effect size (MD=−3.9 sec (95% CI −6.7 to −1.1), t(28)=−2.8, p=0.009, d=−1.04). Finally, the results showed statistically significant between-groups differences in the total time of MIQ-R, also with a large effect size (MD=−10.3 (95% CI −16.2 to −4.4), t(28)=−3.6, p=0.01, d=−1.31). All mental chronometry scores were shorter for the AS than for the CSP patients, whereby the CSP patients took longer to generate mental motor images **(Table 2)**.

### Psychosocial and motor variables

The comparison analysis showed significant between-groups differences in all psychosocial and motor assessed variables except for anxiety levels (MD=−1.2 (95% CI 1.2 to −3.8), t(28)=−0.9, p=0.33) and depressive symptoms (MD=−1.1 (95% CI 1.2 to −3.7), t(28)=−0.89, p=0.38) **(Table 3).**

**Table 3.** Differences between patients and asymptomatic subjects in psychosocial and motor variables.

| **Variables** | **CSP (n=15)** | **AS (n=15)** | **p-value** |
| --- | --- | --- | --- |
| **HADs (Anxiety)** | 6.1 ± 8.3 | 4.8 ± 3.4 | 0.33 |
| **HADs (Depression)** | 4.4 ± 3.8 | 3.2 ± 3.0 | 0.38 |
| **TSK-11** | 25.7 ± 6.3 | 16.6 ± 7.5 | 0.002* |
| **PCS-13** | 13.1 ± 9.5 | 6.3 ± 8.3 | 0.047* |
| **Quick-DASH** | 26.6 ± 13.5 | 13.1 ± 4.9 | <0.001** |
| **CPSS** | 126.6 ± 32.9 | 157.4 ± 31.9 | 0.015* |
| **F-ROM (right) °** | 159.4 ± 26.9 | 174.7 ± 5.8 | 0.041* |
| **F-ROM (left) °** | 154.2 ± 21.7 | 172.8 ± 6.8 | 0.005* |
| **ABD-ROM (right) °** | 151.7 ± 33.8 | 172.3 ± 4.5 | 0.025* |
| **ABD-ROM (left) °** | 147.2 ± 31.3 | 172.8 ± 3.7 | 0.004* |
| **Strength (affected) Kg** | 24.9 ± 14.1 | 36.8 ± 11.2 | 0.017* |

Notes: \* $p < 0.05$ ; \*\* $p < 0.001$ . CSP: Chronic shoulder pain; AS: Asymptomatic subjects; F: Flexion; ABD: Abduction; ROM: Range of movement; °: grades; Kg: Kilograms; HADs: Hospital anxiety and depression scale; TSK-11: Tampa scale of kinesiophobia; PCS-13: Pain catastrophizing scale; DASH: Disabilities of the Arm, Shoulder and Hand; CPSS: Chronic pain self-efficacy scale.

### Correlation analysis

With respect to the correlation analysis in the patients with CSP group, a moderate and positive association was found between the levels of CPSS and the kinesthetic imagining ability (r=0.54, p=0.035). In addition, a moderate and positive association was also found between the strength of the affected upper limb with the total capacity of the imagining questionnaire (r=0.56, p=0.037). Finally, with respect to mental chronometry, a moderate and negative association was found between kinesthetic imagining time and CPSS (r=−0.58, p=0.023), and between visual imagining time and range of motion in flexion of the left upper limb (r=−0.55, p=0.033) **(Table 4)**.

**Table 4.**
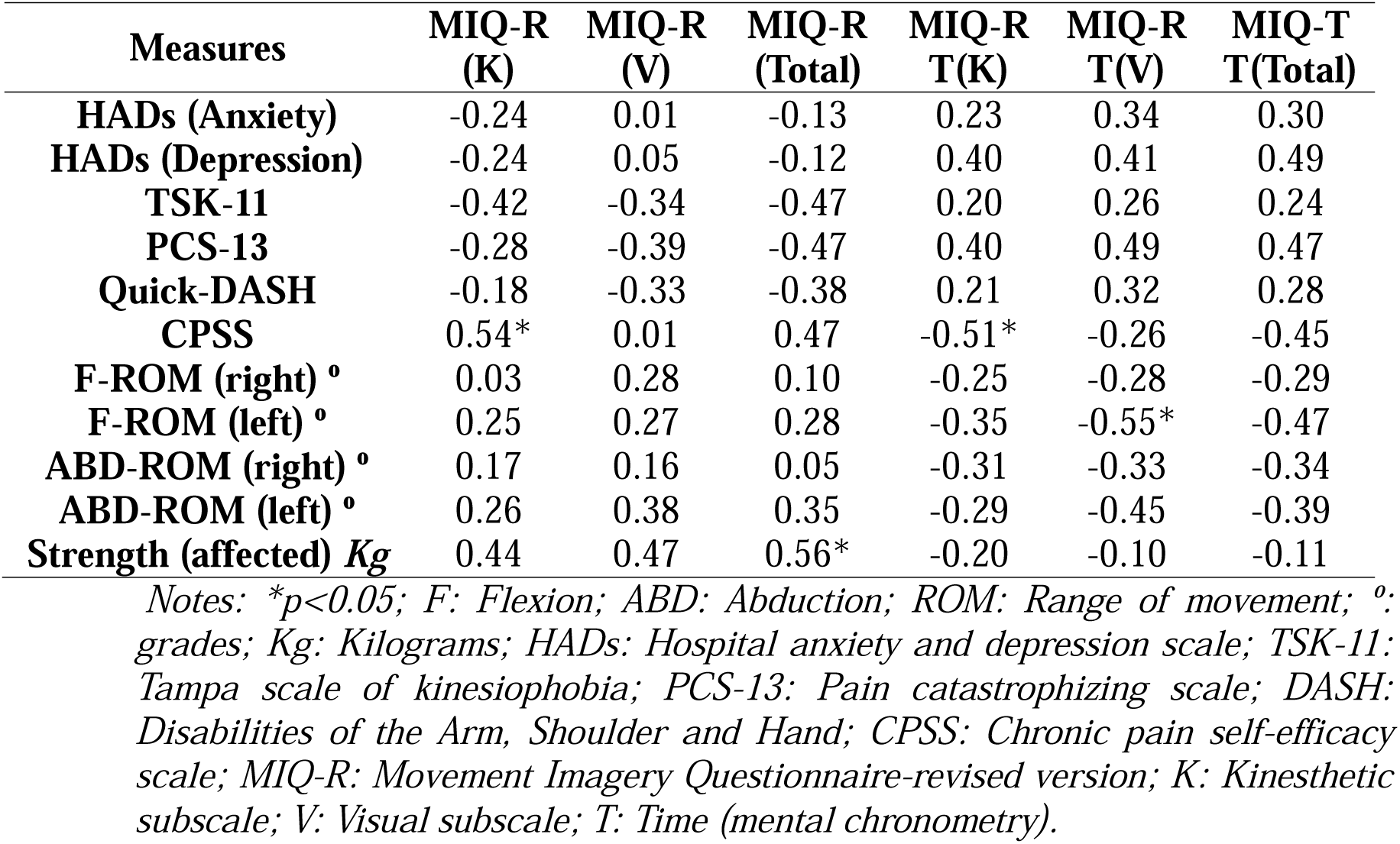
Correlation coefficients between movement imagery and psychosocial and motor variables in patients with chronic shoulder pain.

| Measures | MIQ-R<br>(K) | MIQ-R<br>(V) | MIQ-R<br>(Total) | MIQ-R<br>T(K) | MIQ-R<br>T(V) | MIQ-T<br>T(Total) |
| --- | --- | --- | --- | --- | --- | --- |
| **HADs (Anxiety)** | -0.24 | 0.01 | -0.13 | 0.23 | 0.34 | 0.30 |
| **HADs (Depression)** | -0.24 | 0.05 | -0.12 | 0.40 | 0.41 | 0.49 |
| **TSK-11** | -0.42 | -0.34 | -0.47 | 0.20 | 0.26 | 0.24 |
| **PCS-13** | -0.28 | -0.39 | -0.47 | 0.40 | 0.49 | 0.47 |
| **Quick-DASH** | -0.18 | -0.33 | -0.38 | 0.21 | 0.32 | 0.28 |
| **CPSS** | 0.54* | 0.01 | 0.47 | -0.51* | -0.26 | -0.45 |
| **F-ROM (right) °** | 0.03 | 0.28 | 0.10 | -0.25 | -0.28 | -0.29 |
| **F-ROM (left) °** | 0.25 | 0.27 | 0.28 | -0.35 | -0.55* | -0.47 |
| **ABD-ROM (right) °** | 0.17 | 0.16 | 0.05 | -0.31 | -0.33 | -0.34 |
| **ABD-ROM (left) °** | 0.26 | 0.38 | 0.35 | -0.29 | -0.45 | -0.39 |
| **Strength (affected) Kg** | 0.44 | 0.47 | 0.56* | -0.20 | -0.10 | -0.11 |

*Notes: \*p<0.05; F: Flexion; ABD: Abduction; ROM: Range of movement; °: grades; Kg: Kilograms; HADs: Hospital anxiety and depression scale; TSK-11: Tampa scale of kinesiophobia; PCS-13: Pain catastrophizing scale; DASH: Disabilities of the Arm, Shoulder and Hand; CPSS: Chronic pain self-efficacy scale; MIQ-R: Movement Imagery Questionnaire-revised version; K: Kinesthetic subscale; V: Visual subscale; T: Time (mental chronometry).*

### Simple linear regression analysis

The regression models for criterion variables (MIQ-R kinesthetic subscale and total score of MIQ-R) are presented in **Table 5** and **Table 6**. No regression model was performed for the MIQ-R visual subscale variable (p>0.05). In the first model, the criterion variable MIQ-R kinesthetic subscale was predicted by the CPSS, explaining 24.5% of the variance **(Table 5)**. In the second model, the criterion variable MIQ-R total score was predicted by the strength of affected upper limb, explaining 25.7% of the variance **(Table 6)**.

**Table 5.** Simple linear regression analysis for MIQ-R Kinesthetic Subscale.

| Group | Regression<br>Coefficient<br>(*B*) | Standardized<br>Coefficient<br>($\beta$) | p-value | Overall Model | | |
| --- | --- | --- | --- | --- | --- | --- |
| | | | | $R^2$ | *Adjusted*<br>$R^2$ | *F* |
| CSP |  |  |  | 0.299 | 0.245 | 5.54 |
| **Predictor**<br>**variable** |  |  |  |  |  |  |
| CPSS | 0.077 | 0.54 | 0.035* |  |  |  |

*Notes: \*p<0.05; CSP: Chronic shoulder pain; CPSS: Chronic pain self-efficacy scale; $R^2$ : Determination coefficient.*

**Table 6.**
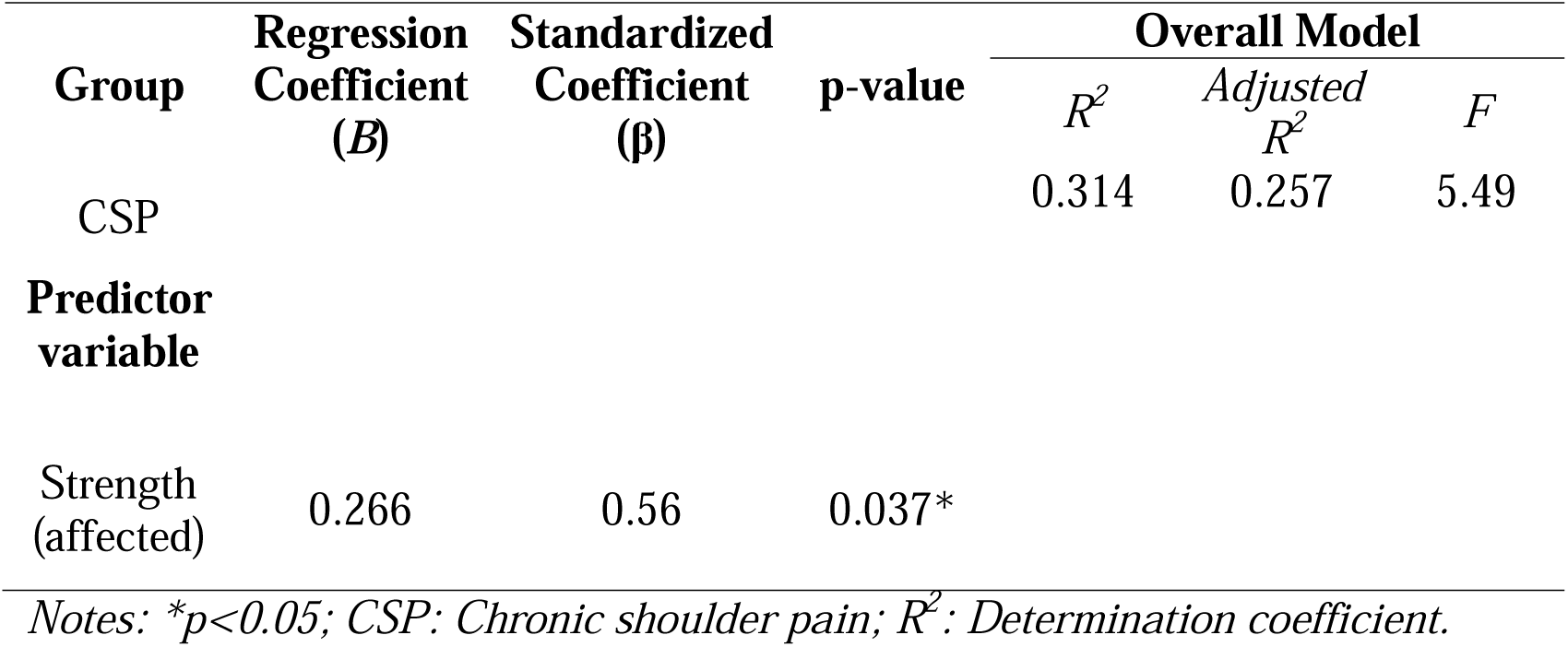
Simple linear regression analysis for MIQ-R total score.

## Discussion

The main aim of the present study was to assess the ability to generate both kinesthetic and visual MI in patients with CSP, compared with AS. In addition, the secondary aim was to assess the influence of psychosocial and motor variables in the MI process.

Regarding the results related to the main objective, it was observed that participants with CSP, compared to AS, had greater difficulties in generating both kinesthetic and visual MI and needed more time to execute the mental tasks. One of the causes that may explain these results is the presence of alterations at the cortical level in patients with CSP. In relation to these cortical changes, one of the possible alterations involved is the alteration of functional connections associated with motor planning and execution due to increased activity of default mode networks in response to pain (Tagliazucchi et al., 2010). On the other hand, the other possible cortical alteration involved is the distortion of the affected area in body representation at the cortical level due to the maintenance of pain (Moseley et al., 2006). Therefore, the difference in participants’ ability to produce motor imagery may be directly related to both changes.

These results are in line with the findings obtained by La Touche et al. 2019 in patients with chronic low back pain. La Touche et al. 2019 saw that patients with chronic low back pain compared to asymptomatic subjects had a reduced ability to generate MI, both visual and kinaesthetic, as well as the need for more time to perform MI (La Touche et al., 2019).

As for the secondary objectives, firstly, in terms of psychosocial variables, it is worth mentioning that the results indicate the existence of a relationship between the level of chronic pain self-efficacy, measured by the CPSS, and the kinesthetic imagination. Our results indicate that there is a moderate and positive relationship between the levels obtained in the CSSP and the ability to generate kinesthetic MI. It was also found that the higher the CSSP score, the less time was needed to generate kinesthetic motor images. All this may be mainly because higher self-efficacy is related to lower disability which may translate into a higher capacity to generate kinesthetic MI (La Touche et al., 2019).

On the other hand, regarding the motor variables measured, it was observed that both the ROM and the strength of the affected upper limb were related to the ability to generate MI. On one side, the results indicated that the longer the flexion ROM of the left upper limb, the shorter the time required to generate visual MI. On the other side, it was found that the strength of the affected upper limb was positively associated with the MIQ-R total score, meaning that the greater the strength of the affected upper limb, the higher the MIQ-R total score obtained. We can therefore conclude that the better the patient’s motor status, the greater the ability to generate MI.

Finally, it is important to note that at the clinical level, the existence of impairments in the ability of patients with CSP to generate both visual and kinesthetic MI may directly affect the use of MI as a treatment tool, either in isolation or as part of a graded motor imagery procedure (Araya-Quintanilla et al., 2020). Therefore, it would be of great interest to analyze the ability of patients with CSP to generate motor imagery, both visual and kinesthetic, prior to the use of MI for therapeutic purposes in order not to overlook the possible existence of MI disturbances.

## Limitations

There are various limitations on this study. Firstly, the cross-sectional study design implies that the results should be considered cautiously because it difficult to determine a cause-and-effect link. Secondly, it might be interesting in the future to evaluate the medication taken by patients as there are certain medicines that can affect the generation of MI. Finally, the complex process of producing MI was evaluated using a mental chronometry task and a self-report instrument. Even though we think these instruments have acceptable reliability, it could be interesting to examine the habit of employing neural functional images to generate motor images in future investigations.

## Conclusions

Based on the obtained results, it seems that patients with CSP have greater difficulty with generating both kinesthetic and visual motor images compared to AS. In addition, patients with CSP needed more time to perform the motor mental tasks compared to AS. Finally, it was found that psychosocial state and motor condition may influence the imagining ability of patients with CSP where greater self-efficacy levels and muscle strength in patients with CSP are correlated with more ability to create motor images. Further research is needed to improve our understanding and to be able to establish clinical implications for the results obtained in this study.

## Data Availability

All data produced in the present study are available upon reasonable request to the authors

